# Prenatal extreme heat exposure and neonatal mortality in Bangladesh, 2015 to 2025: a national birth-history study

**DOI:** 10.64898/2026.10.23.26366352

**Authors:** S. M. Ashraf Ul Hoque, Md Khairul Islam, Mohammad Khalid Hasan, Md Mohiuddin Sharif, S M Raysul Haque, Kamran ul Baset

## Abstract

**Background:** Heat in pregnancy is linked to preterm birth and stillbirth, but its link with neonatal death in South Asia is unclear. We estimated this association in Bangladesh and separated the role of prenatal heat from that of the season in which a child is born.

**Methods:** We included 46,752 live births (February 2015 to May 2025) from the Bangladesh Multiple Indicator Cluster Survey 2025 birth history. Each birth was linked to daily maximum temperature from the ERA5 reanalysis at the location of its survey cluster. An extreme heat day was a day on which the maximum temperature reached the cluster’s 95th percentile of daily maximum temperature for 2014 to 2024. Because gestational age was not recorded, we counted extreme heat days in estimated trimesters, dated back 280 days from birth. Weighted Poisson regression with fixed effects for district, division by calendar month of birth, and year of birth compared births within the same place, season and year. Extreme heat 28 to 55 days after birth, which cannot cause a neonatal death, served as a negative control exposure.

**Results:** There were 1,039 neonatal deaths (22.7 per 1,000 live births, 95% CI 21.1 to 24.4). Mortality was highest for births in December to February (26.5 per 1,000, 23.0 to 29.9) and lowest in September to November (17.3, 14.5 to 20.0). Pregnancies had a mean of 13.5 extreme heat days (95% CI 13.4 to 13.6), rising from 4.0 for births in 2018 to 29.5 for births in 2024. The primary estimate was for the estimated third trimester. Per 5 extreme heat days, its adjusted rate ratio was 1.06 (0.98 to 1.15), a risk difference of 1.4 per 1,000 births (−0.6 to 3.3). The rate ratio was 1.04 (0.98 to 1.09) for the whole pregnancy and 0.92 (0.77 to 1.09) for the last 4 weeks of pregnancy. The negative control window showed no clear association (0.92, 0.79 to 1.08). Third trimester estimates were similar with MERRA-2 temperature (1.04, 0.97 to 1.12) and with a fixed 1991 to 2020 threshold (1.07, 1.00 to 1.14). Without control for calendar month of birth, each 1 °C higher mean third trimester maximum temperature was associated with lower mortality (rate ratio 0.96, 0.93 to 0.99).

**Conclusions:** In this national birth-history analysis, neonatal mortality varied by season of birth and was highest for births in December to February. When births were compared within the same district, season and year, estimated prenatal extreme heat was not clearly associated with neonatal death. The confidence intervals did not exclude modest increases in risk, and the study had 80% power only for a rate ratio of 1.12 per 5 days or larger. Both gestational age and heat exposure were estimated with error. Studies of prenatal heat and newborn survival should control for season of birth, because season can hide or create an apparent effect of heat.

## Introduction

Bangladesh had a neonatal mortality rate of 22 per 1,000 live births in the five years before 2025 [1]. The Sustainable Development Goals ask every country to reach 12 per 1,000 or less by 2030 [2]. Climate change adds a new pressure on this target [3]. Global mean surface temperature reached a record 1.45 °C above the pre-industrial baseline in 2023 [4].

Heat in pregnancy is linked to adverse birth outcomes. In meta-analyses, the odds of preterm birth rose by about 4 to 5% per 1 °C of heat exposure, and stillbirth risk also rose with temperature [5,6]. Proposed pathways include dehydration, reduced placental blood flow and an inflammatory response that may trigger labour [7]. Among pregnant farmers in The Gambia, heat stress was associated with fetal strain [8]. Preterm birth complications and intrapartum events are leading causes of neonatal death [9], so heat in pregnancy could also raise neonatal mortality.

Fewer studies have examined neonatal death, and their results differ. A study of 29 low-income and middle-income countries, including Bangladesh, linked neonatal deaths to temperature in the days before death [3]. It attributed 1.5% of neonatal deaths to heat and 2.9% to cold. Studies of heat in pregnancy and neonatal death come mainly from sub-Saharan Africa [10] or pool many countries without describing control for season of birth [11]. A case-crossover study in East Africa examined temperature after birth and gave imprecise pooled estimates [12]. Season matters because births in different months differ in heat exposure and in other risks, such as cold after birth [13,14]. Studies of prenatal heat that include Bangladesh have examined extreme low birthweight and stunting, not neonatal death [15,16]. No national study has asked whether heat during pregnancy, as distinct from the season of birth, is associated with neonatal death in Bangladesh.

The Multiple Indicator Cluster Survey (MICS) 2025 makes this question answerable. It recorded the day of birth of every child in a full birth history and released displaced locations for all survey clusters [1,17]. Because the birth histories extend to mid-2025, they also cover the recent hot years.

Our overall objective was to estimate whether extreme heat during pregnancy is associated with neonatal death in Bangladesh. Our first aim was to describe how often pregnancies were exposed to extreme heat, and how exposure and neonatal mortality varied by month of birth. Our second aim was to estimate the association between extreme heat in estimated windows of pregnancy and neonatal death, comparing births within the same place, season and year. We used a negative control window after the neonatal period to look for residual confounding. Our third, exploratory aim was to examine whether the association differed between the poorest 40% of households and the rest, and between urban and rural areas.

## Methods

### Data source and design

MICS 2025 was a nationally representative household survey by the Bangladesh Bureau of Statistics with the United Nations Children’s Fund (UNICEF) [1]. It used a two-stage stratified cluster sample covering all 64 districts. Fieldwork ran from 24 February to 30 June 2025. Forty-three teams, each with five trained female interviewers, a measurer, a phlebotomist and a supervisor, collected data on tablets by computer-assisted personal interviewing. The questionnaires followed the MICS7 standard and were translated into Bangla and pretested. Supervisors monitored fieldwork daily and re-interviewed one household per cluster, and field check tables were reviewed weekly. The household response rate was 99.6%, and 64,405 eligible women aged 15 to 49 years (95.8%) were interviewed. Ever-married women gave a full birth history, with the day, month and year of each live birth and age at death of children who died. Never-married women were not asked the birth history. Births are therefore observed only through women alive, resident and aged 15 to 49 years at the 2025 interview.

### Study population

We included live births 1 to 120 months before the interview month (n=46,858). We used 10 years of births to increase the number of deaths; an analysis restricted to births in the last 5 years was a sensitivity analysis. We excluded 106 births within 28 days before the interview, because their neonatal period was incomplete. The analytic sample was 46,752 births to 32,526 women in 3,149 clusters (S1 Fig). Day or month of birth was unknown for 102 births; we set the day to 15 and took the month from the survey’s imputed date. Eleven births lacked mother’s education and no other model variable was missing. Because missing data were so few, adjusted models used complete cases (n=46,741).

### Outcome

Neonatal death was defined as death at age 0 completed months, the definition used in the MICS report [1]. As a check of coding, the weighted neonatal mortality for births 1 to 60 months before the interview was 21.9 per 1,000, against 22 in the report [1].

### Heat exposure

MICS displaced each cluster location by up to 2 km in urban and 5 km in rural areas, within the same upazila (subdistrict) [17]. Daily maximum temperature and daily mean wet bulb temperature for 1 January 2014 to 31 July 2025 came from ERA5 [18]. ERA5 is the fifth-generation global reanalysis of the European Centre for Medium-Range Weather Forecasts. We obtained it through the Open-Meteo archive interface [19] with its ERA5 model setting. Open-Meteo aggregates hourly data into local-time daily values and supplies wet bulb temperature as a derived variable, without documenting its formula. We requested the 98 points of a 0.5° grid of points covering Bangladesh, so no cluster location left the study computer. For each point, the service returned the value of the native ERA5 grid cell, which has a resolution of 0.25°, using its default cell selection. We interpolated these values bilinearly from the four nearest points to each cluster point.

An extreme heat day was a day with maximum temperature at or above the cluster’s 95th percentile of all days in 2014 to 2024. A local threshold reflects the heat to which each population is adapted. Using the study years as reference defines extremes against the climate people lived in, while birth-year fixed effects absorb year-level shifts. Days in January to July 2025 were classified with the same 2014 to 2024 threshold. Thresholds ranged from 30.8 °C to 37.4 °C across clusters (median 35.4 °C). As a sensitivity analysis, we also used a fixed 1991 to 2020 ERA5 reference period, the current climate normal.

Gestational age was not recorded. We therefore dated the last menstrual period as 280 days before birth and counted extreme heat days in estimated trimesters. These were the first (280 to 190 days before birth), the second (189 to 92 days) and the third (91 to 1 day). These are calendar windows counted back from birth, not biological trimesters. The first and third windows span 91 days and the second spans 98 days, so counts per 5 days are not equally scaled across windows. We also counted extreme heat days over the whole pregnancy and in the last 4 weeks before birth. The negative control window was days 28 to 55 after birth. Heat in this window cannot cause a neonatal death, but it shares the seasonal and spatial pattern of the other windows [20]. As an exploratory measure, we counted extreme heat days from birth to the day of death, or to day 27 for survivors, as a rate per 28 days. This measure therefore includes no heat after death.

We also defined a temperature anomaly. For each cluster and day, it was the daily maximum temperature minus the cluster’s 2014 to 2024 mean for that calendar day, smoothed over 31 days. We averaged it over each window.

We rebuilt every exposure from a second reanalysis, the Modern-Era Retrospective analysis for Research and Applications, Version 2 (MERRA-2) [21]. We obtained it from the Prediction Of Worldwide Energy Resources (POWER) service of the National Aeronautics and Space Administration (NASA), on its 0.5° by 0.625° grid [22]. Across 13.3 million cluster-days, daily maximum temperatures in the two products correlated well (r=0.86; MERRA-2 minus ERA5 −0.2 °C). Agreement on extreme heat days was moderate (kappa 0.53; S1 Table). ERA5 ranked 2023 and 2024 as the years with the most extreme heat days; MERRA-2 did not. MERRA-2 maximum temperature also drifted downward, from 31.0 °C in 1991 to 2000 to 29.7 °C in 2015 to 2024 over interior Bangladesh. We therefore used ERA5 for the main analysis.

### Covariates

Confounders were selected a priori, not by statistical testing. We chose risk factors for neonatal death that could also differ by timing or place of birth. These were sex, multiple birth, birth order and preceding interval, mother’s age at birth, mother’s education, household wealth quintile and urban or rural residence. The MICS wealth index is a principal components score of household assets and housing characteristics, grouped into quintiles [1]. Wealth quintile and mother’s education were measured at the interview, not at birth.

### Statistical analysis

For aim 1, we estimated the mean number of extreme heat days per pregnancy and the share of births with at least 1, 5, 10 and 20 days. We also counted mean extreme heat days per calendar year across survey clusters. We also mapped district means for births in 2015 to 2019 and 2020 to 2025, using geoBoundaries outlines [23]. We estimated neonatal mortality and exposure by calendar month of birth, and tested equal mortality across months with a design-based Wald test.

For aim 2, we fitted Poisson regression models for neonatal death, with women’s sample weights and standard errors clustered by primary sampling unit. Births are observed through interviewed women, so each birth carries its mother’s weight, as in MICS mortality estimates. Every woman’s births lay in one primary sampling unit, so this clustering also covers correlation between siblings. Exposure was entered per 5 extreme heat days. Fixed effects were district (64 levels), division by calendar month of birth (96 levels), and year of birth (11 levels). With 1,039 deaths, the model has about 6 events per estimated parameter. We therefore checked that estimates were stable as fixed effects were added (S3 Table). The heat estimate therefore compares births with more or fewer extreme heat days within the same district, calendar month within division, and year of birth. These fixed effects do not remove changes over time within districts. The model is written out in S1 Text.

The main model included the three estimated trimesters together, with all covariates and no exposure after birth. No protocol was registered. We designated the third trimester term as the primary estimate because it is the window closest to birth. This designation was made during the analysis, after an earlier version of the model, so it is a design choice and not a pre-registered one.

All other estimates were secondary. The three trimester counts were weakly correlated (Spearman −0.37 to −0.01; S2 Table). A second model split the third trimester into the last 4 weeks and the earlier weeks. A third model used the whole pregnancy count alone. A fourth added the negative control window to the main model. A fifth added the exploratory neonatal exposure to the main model. Risk differences per 1,000 births came from linear probability models with the same terms. Dose-response was examined with categories of third trimester exposure (0, 1 to 4, 5 to 9 and 10 or more days), shown for display. As the main check of non-linearity, we fitted third trimester exposure as a restricted cubic spline. Its three knots sat at the 10th, 50th and 90th percentiles of non-zero values (1, 4 and 20 days). Non-linearity was tested with a Wald test. From the standard error of the primary estimate, we calculated the smallest rate ratio per 5 days detectable with 80% power at a two-sided 5% level.

For aim 3, which was exploratory, we added an interaction between third trimester exposure and the poorest 40% of households. In a separate model, we added an interaction with rural residence.

Five sensitivity analyses addressed named threats. First, mothers who moved during pregnancy were assigned the heat of their current home, not of where they lived while pregnant. We therefore restricted to mothers living in the same place since before the last menstrual period. Second, dry heat may not capture humid heat stress, so we counted days of mean wet bulb temperature at or above the cluster’s 95th percentile. Third, recall of birth dates and deaths is weaker for older births, so we restricted to births in the 5 years before the survey. Fourth, the two reanalyses disagreed on many extreme days, so we refitted all models with MERRA-2 exposures (S1 Table). Fifth, the 2014 to 2024 reference overlaps the exposure years, so we used the fixed 1991 to 2020 threshold.

To show how the design affects estimates, we refitted single-window models with fewer fixed effects, from covariates and division only to the main set (S3 Table). Because windows are calendar proxies, we also used single windows of 28 to 120 days before birth and an assumed 259-day pregnancy (S4 Table). We compared variance estimators, including a fully design-based model with strata (S4 Table). Because exposure varies mainly between years and regions, we also clustered standard errors by district and birth year, division and birth year, and birth year alone (S4 Table). A wild cluster score bootstrap with Webb weights at these levels gave p values and intervals, found by inverting the test (S4 Table). Further checks used a 90th percentile threshold, temperature anomalies, district by month fixed effects, district-specific linear trends and division by year fixed effects. Other checks changed the outcome to early neonatal death or to death on days 0 to 27, and excluded births with imputed dates (S4 Table). Analyses used R 4.6.1 with the fixest and survey packages [24,25]. We report 95% confidence intervals (CIs) and exact two-sided p values. The study follows the Strengthening the Reporting of Observational Studies in Epidemiology (STROBE) guideline [26].

### Ethics

The icddr,b Ethical Review Committee approved MICS 2025 (PR-24113) [1]. Respondents gave verbal informed consent. This secondary analysis used anonymised public data and needed no further approval.

## Results

The 46,752 births included 1,039 neonatal deaths, a weighted rate of 22.7 per 1,000 (95% CI 21.1 to 24.4; Table 1). Pregnancies had a mean of 13.5 extreme heat days (95% CI 13.4 to 13.6; median 9, interquartile range 4 to 21; S2 Table). Overall, 95.3% of births had at least one extreme heat day in pregnancy and 48.9% had 10 or more. In the estimated third trimester, 53.0% had at least one and 25.2% had 5 or more (S2 Table).

**Table 1.**
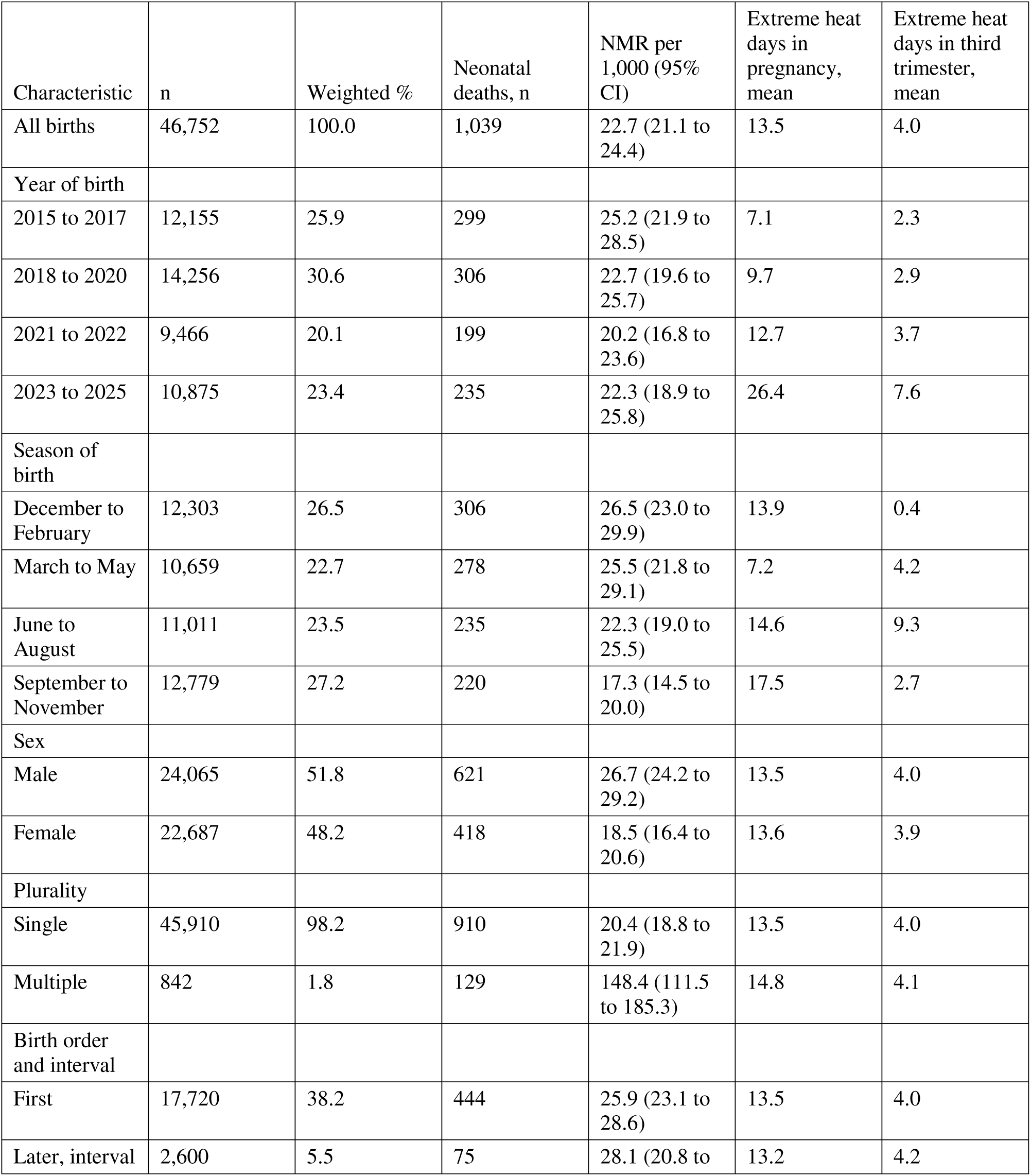
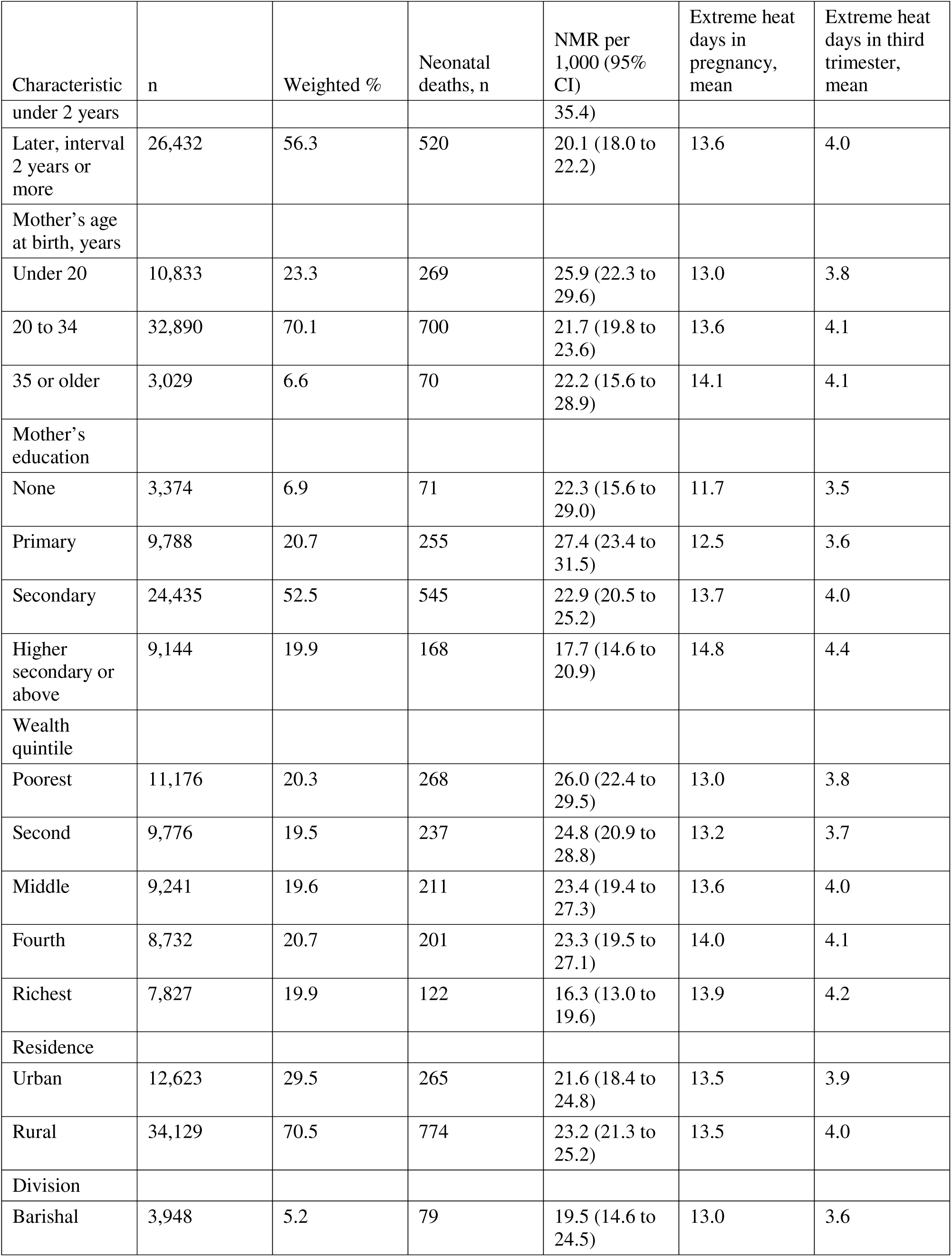
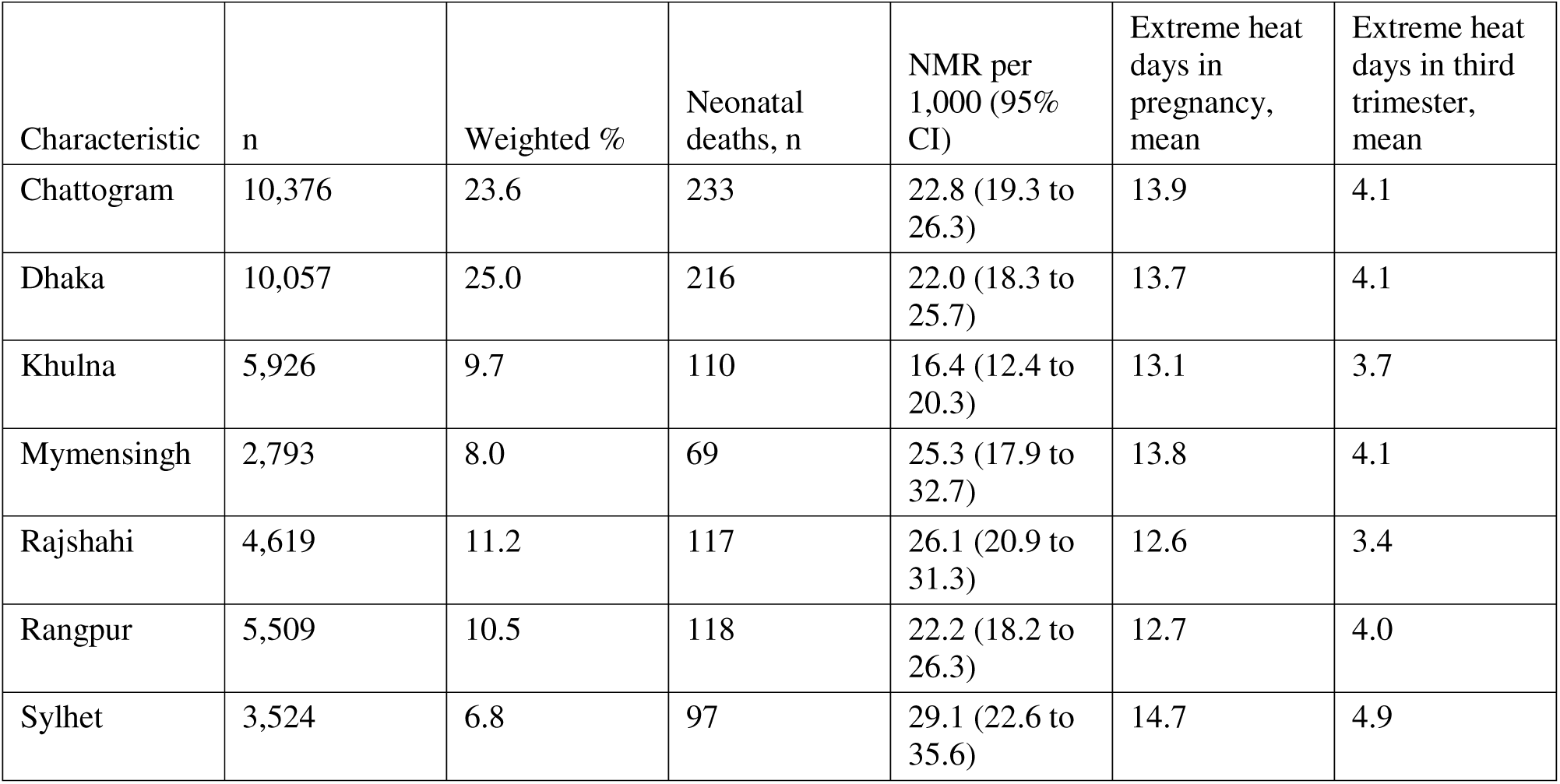
Live births, neonatal mortality and extreme heat in pregnancy, Bangladesh MICS 2025, births 2015 to 2025. n is unweighted. Percentages, rates and means use survey weights; 95% CIs use the survey design. An extreme heat day had a daily maximum temperature at or above the cluster’s 95th percentile for 2014 to 2024 (ERA5). NMR, neonatal mortality rate. Eleven births lack mother’s education.

At survey clusters, the most extreme heat days fell in 2024 (37.3 per cluster) and 2023 (35.6), against 5.4 in 2018 (Fig 1A). The fixed 1991 to 2020 threshold was on average 0.41 °C lower. Under it, clusters had 46.6 extreme heat days in 2024 and 45.0 in 2023, against 11.7 in 2018 (Fig 1A; S1 Table). Exposure per pregnancy rose from a mean of 4.0 days for births in 2018 to 24.3 in 2023 and 29.5 in 2024 (Fig 1B; S2 Table). Every district had higher exposure for births in 2020 to 2025 than in 2015 to 2019 (Fig 2). District means ranged from 5.6 to 12.4 days in the earlier period and from 13.3 to 22.0 in the later one (Fig 2B). Local thresholds ranged from 33.2 °C to 36.9 °C across districts (Fig 2A).

**Fig 1.**
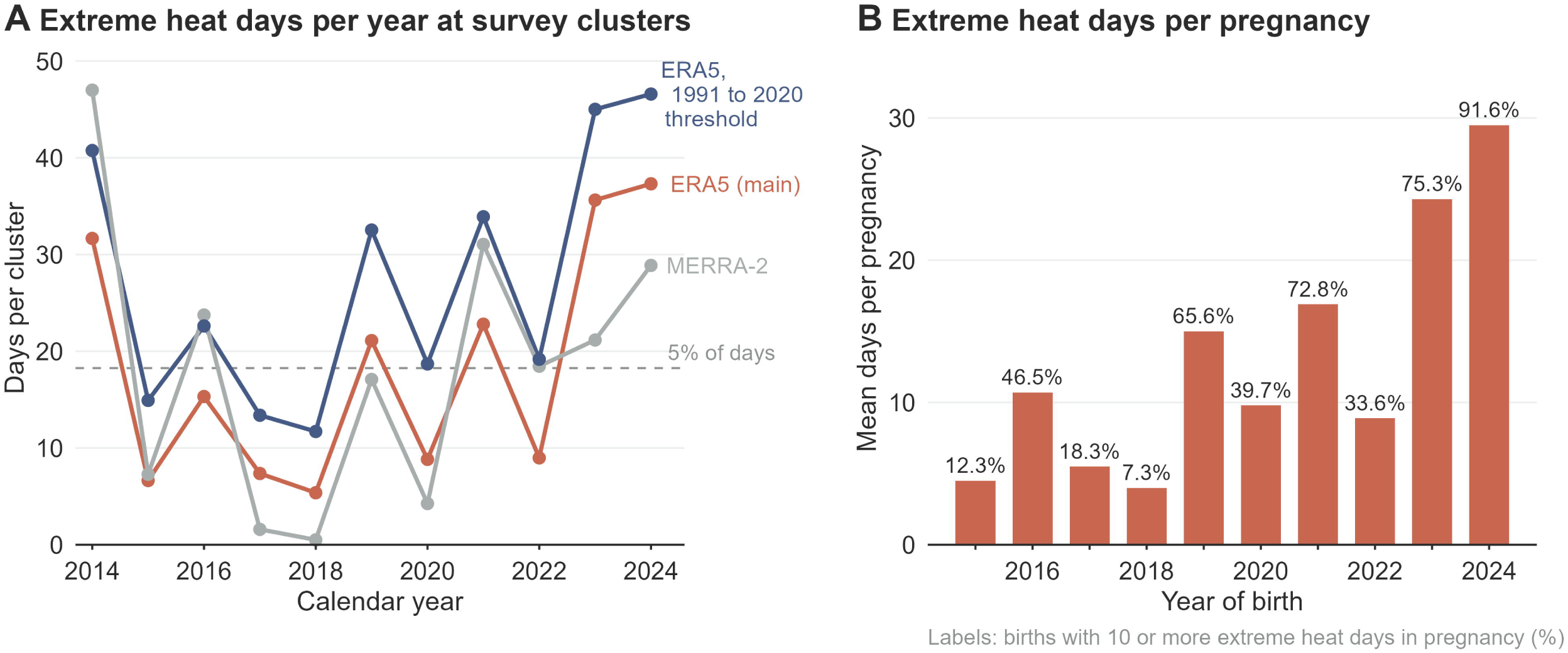
Extreme heat days at survey clusters and during pregnancy, Bangladesh, 2014 to 2024. (A) Mean extreme heat days per calendar year across 3,149 survey clusters: ERA5 and MERRA-2 with the 2014 to 2024 threshold, and ERA5 with a fixed 1991 to 2020 threshold. The dashed line marks 5% of days, the average expected under the main threshold. (B) Weighted mean extreme heat days per pregnancy by year of birth (ERA5, main threshold). Labels give the share of births with 10 or more such days.

**Fig 2.**
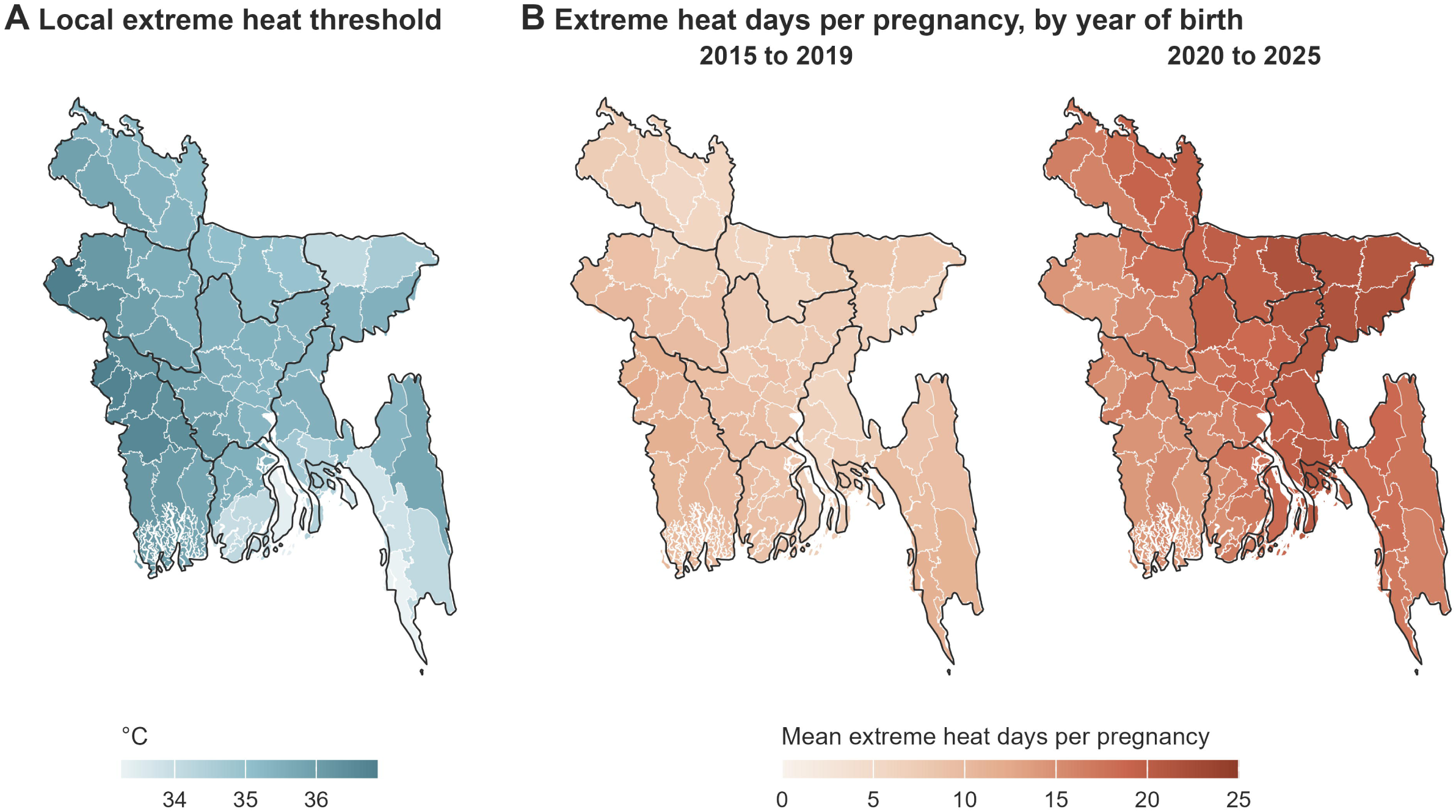
Local heat threshold and extreme heat in pregnancy by district, Bangladesh, births 2015 to 2025. (A) Mean cluster 95th percentile of daily maximum temperature, 2014 to 2024 (ERA5). (B) Weighted mean extreme heat days per pregnancy for births in 2015 to 2019 and 2020 to 2025, on one shared scale. Thick lines are division boundaries. Boundaries from geoBoundaries [23].

Neonatal mortality differed by calendar month of birth (p=0.008; Fig 3). It was highest among births in December to February (26.5 per 1,000, 95% CI 23.0 to 29.9) and lowest in September to November (17.3, 14.5 to 20.0). Third trimester heat exposure followed a different seasonal pattern from mortality. Births in June and July had a mean of 12.8 and 10.6 third trimester extreme heat days, while births in January to March had almost none.

**Fig 3.**
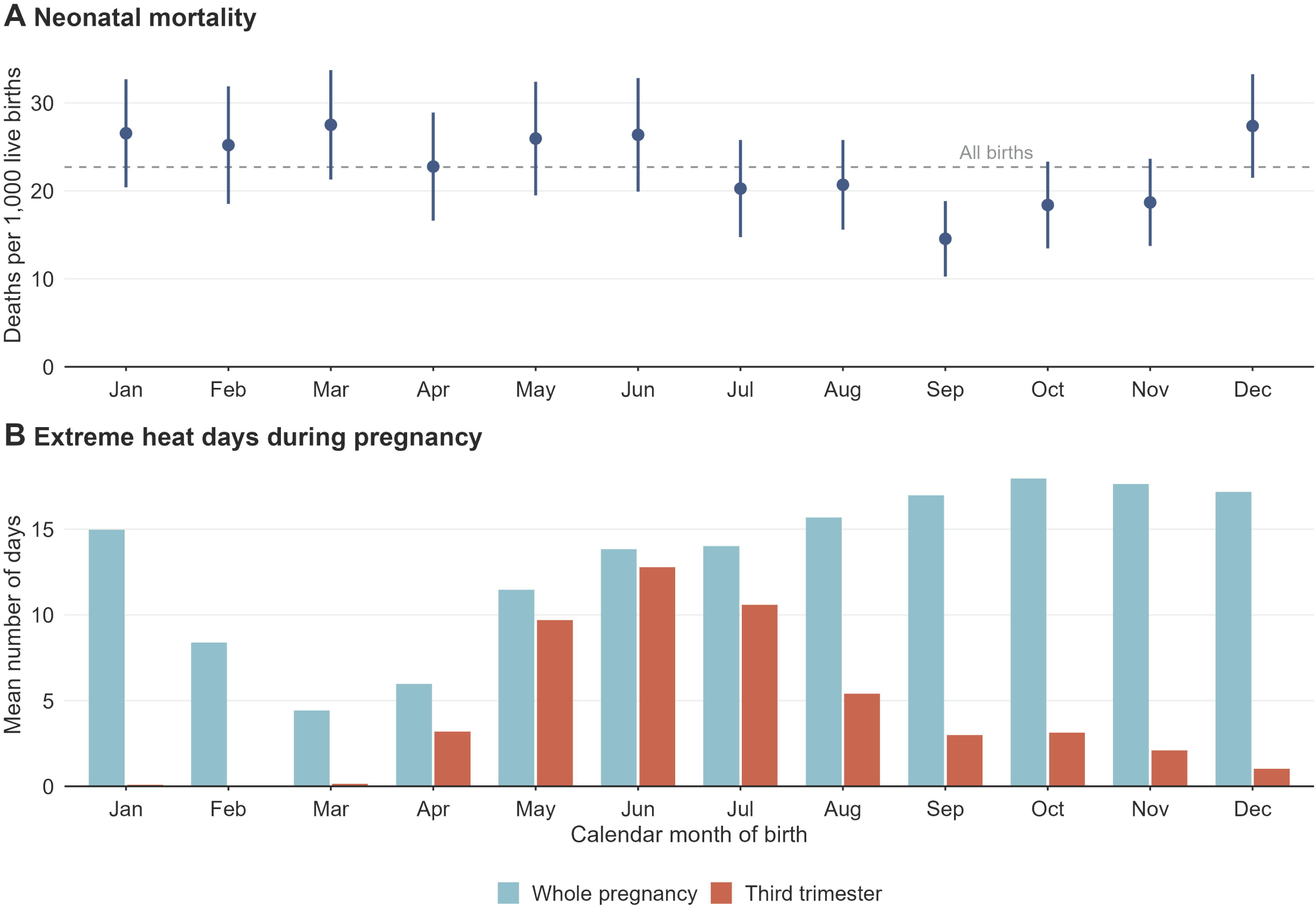
Neonatal mortality and extreme heat in pregnancy by calendar month of birth, Bangladesh, births 2015 to 2025. (A) Weighted neonatal mortality per 1,000 live births with 95% CIs; the dashed line is the rate for all births. (B) Mean extreme heat days in the whole pregnancy and in the estimated third trimester (ERA5). n=46,752 births.

After the fixed effects for place, season and year were removed, 43% of the variance in the third trimester count remained (standard deviation 4.5 days; S2 Table). We found no clear evidence of an association between extreme heat in any estimated window and neonatal death (Table 2; Fig 4A). Per 5 extreme heat days, the adjusted rate ratio was 1.02 (95% CI 0.95 to 1.09) for the first trimester and 1.04 (0.96 to 1.12) for the second. It was 1.06 (0.98 to 1.15) for the third trimester, a risk difference of 1.4 per 1,000 births (−0.6 to 3.3). The smallest rate ratio per 5 days that the study could detect with 80% power was 1.12.

**Fig 4.**
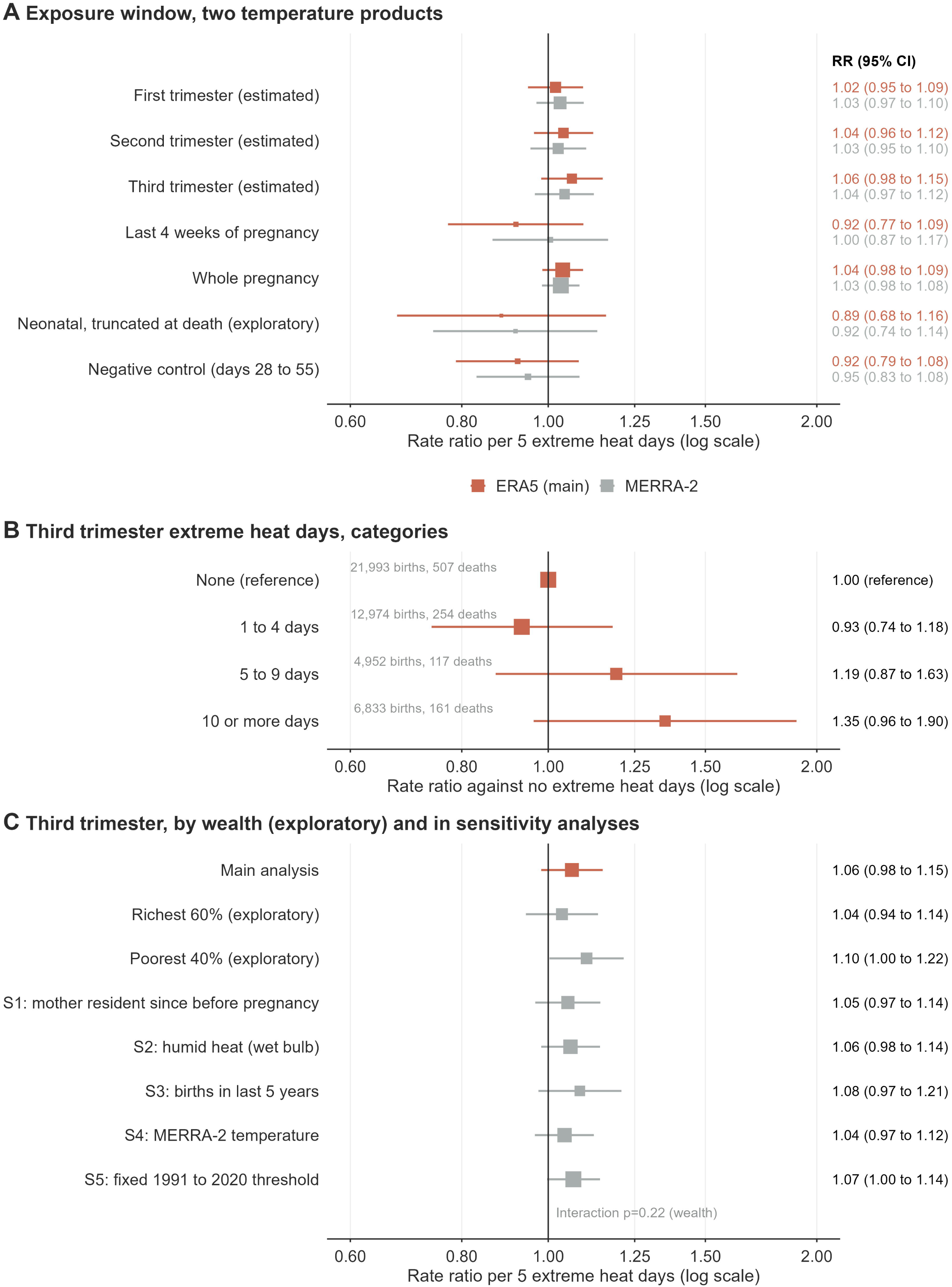
Rate ratios for neonatal death and extreme heat in pregnancy. (A) Per 5 extreme heat days by estimated window, with ERA5 (main) and MERRA-2 exposures. The neonatal term counts heat only up to death and is exploratory; the negative control window falls after the neonatal period. (B) Third trimester categories against no extreme heat days, with unweighted births and deaths. (C) Third trimester, by household wealth (exploratory) and in five sensitivity analyses. Squares show adjusted rate ratios, sized by statistical information; lines show 95% CIs. All models include fixed effects for district, division by calendar month of birth and year of birth. RR=rate ratio.

**Table 2.**
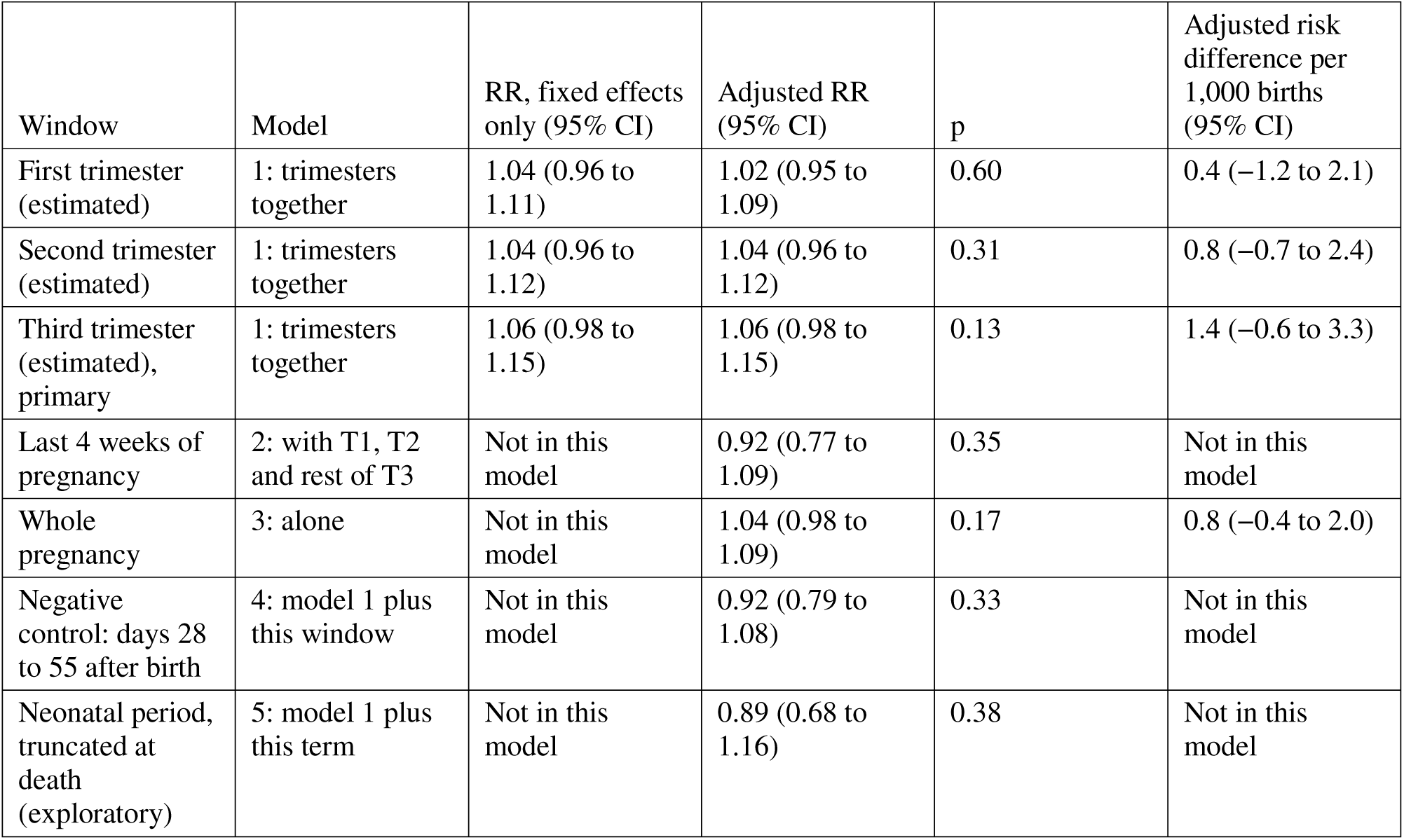
Neonatal death per 5 extreme heat days, by estimated exposure window. RR, rate ratio from weighted Poisson regression. All models include fixed effects for district, division by calendar month of birth and year of birth. “Not in this model” marks a term or estimate not produced by that model. Model 1 holds the three estimated trimesters together and is the main model; the fixed effects only column omits covariates. Adjusted models add sex, multiple birth, birth order and interval, mother’s age, education, wealth quintile and residence. Risk differences come from linear probability models with the same terms. The exploratory neonatal term counts extreme heat days from birth to death or day 27, per 28 days. Standard errors are clustered by primary sampling unit. n=46,741 births and 1,039 deaths.

The adjusted rate ratio was 0.92 (0.77 to 1.09) for the last 4 weeks of pregnancy. For the whole pregnancy it was 1.04 (0.98 to 1.09), a risk difference of 0.8 per 1,000 births (−0.4 to 2.0) per 5 days. The negative control window showed no clear association (0.92, 0.79 to 1.08). It follows the neonatal period, so the two counts were correlated (Spearman 0.43), but it was not correlated with the third trimester count (−0.02; S2 Table). The exploratory neonatal exposure, truncated at death, had a ratio of 0.89 (0.68 to 1.16). Third trimester exposure was also grouped into categories. Births with 10 or more extreme heat days had a ratio of 1.35 (0.96 to 1.90) against births with none (Fig 4B; S4 Table). Births with 1 to 4 days had a ratio of 0.93 (0.74 to 1.18). The spline gave no evidence of departure from linearity (p=0.21). Its curve rose to 1.34 (0.98 to 1.83) at 10 days and then flattened, with intervals including 1 throughout (S2 Fig).

The choice of fixed effects changed the estimates for mean third trimester temperature (Fig 5; S3 Table). With only division fixed effects and covariates, each 1 °C higher mean third trimester maximum temperature had a ratio of 0.96 (0.93 to 0.99; p=0.003). With control for calendar month of birth, as in the main specification, the ratio was 1.03 (0.94 to 1.12).

**Fig 5.**
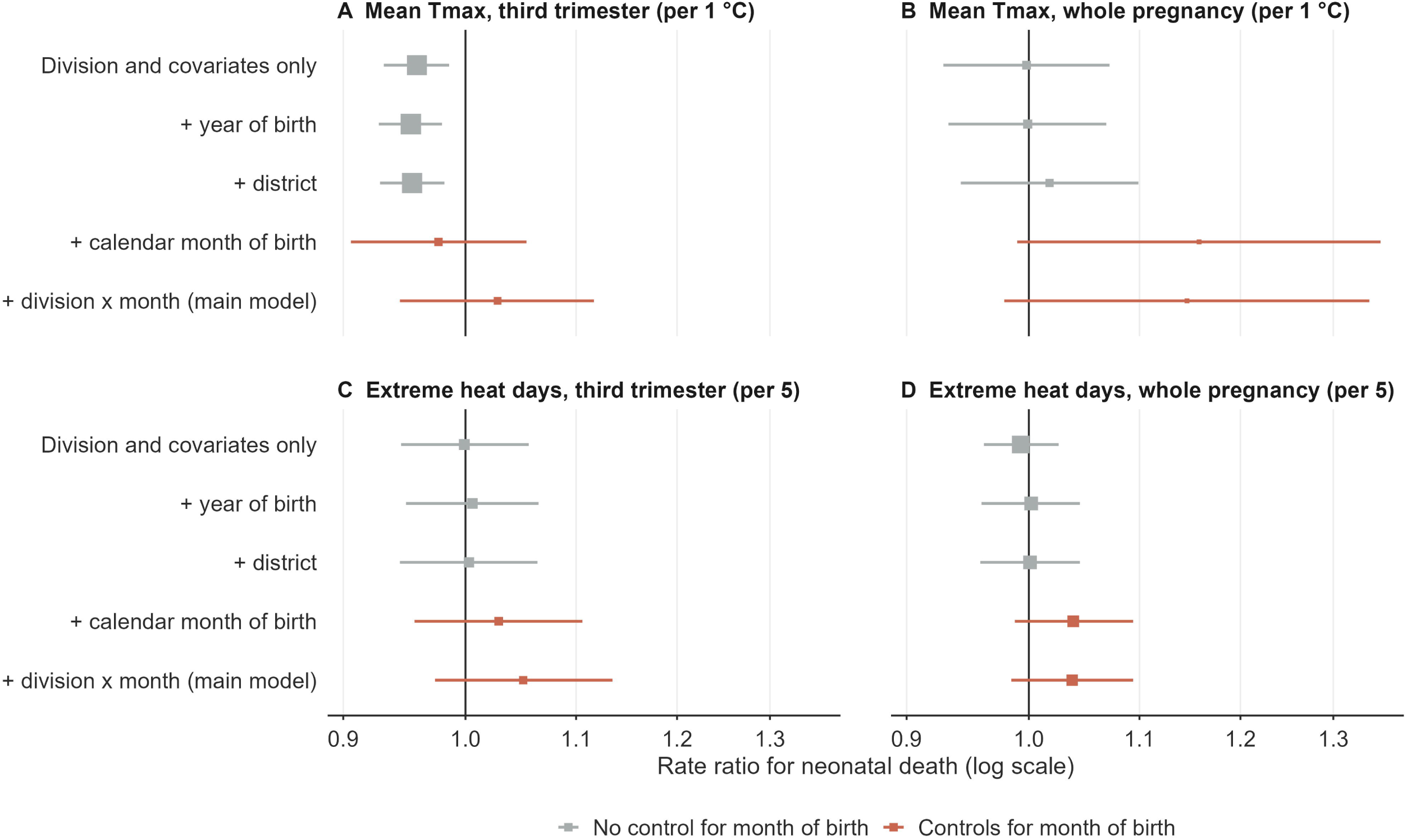
Estimates for heat in pregnancy as fixed effects are added. Rate ratios for neonatal death with 95% CIs, from models with one exposure each and all covariates. Rows add fixed effects in turn; terracotta rows control for calendar month of birth. (A, B) Mean daily maximum temperature (Tmax) per 1 °C. (C, D) Extreme heat days per 5 days. ERA5 exposures. n=46,741 births.

In the exploratory wealth analysis, the third trimester ratio was 1.10 (1.00 to 1.22) in the poorest 40% of households and 1.04 (0.94 to 1.14) in the rest. The interaction p value was 0.22 (Fig 4C). The ratio was 1.07 (0.96 to 1.20) in urban areas and 1.06 (0.97 to 1.15) in rural areas (interaction p=0.83; S4 Table).

All five sensitivity analyses gave third trimester estimates between 1.04 and 1.08 (Fig 4C). Restricted to mothers living in the same place since before pregnancy (n=43,766), the ratio was 1.05 (0.97 to 1.14). When extreme heat days were defined by wet bulb temperature to capture humid heat, it was 1.06 (0.98 to 1.14). For births in the 5 years before the survey (n=23,645), it was 1.08 (0.97 to 1.21). With MERRA-2 exposures, it was 1.04 (0.97 to 1.12), and the negative control showed no clear association (0.95, 0.83 to 1.08; S1 Table). With the fixed 1991 to 2020 threshold, it was 1.07 (1.00 to 1.14).

Alternative calendar windows gave ratios from 0.92 (0.78 to 1.09) for the last 28 days to 1.04 (0.97 to 1.11) for the last 120 days (S4 Table). With an assumed 259-day pregnancy, the third trimester ratio was 1.05 (0.95 to 1.15). Every variance estimator gave the same third trimester interval within 0.01, including the fully design-based model (1.06, 0.98 to 1.15; S4 Table). The unweighted model gave the same estimate (1.06, 0.99 to 1.14; S4 Table). Clustering by district and birth year gave 1.06 (0.98 to 1.15), and clustering by division and birth year gave 1.06 (0.99 to 1.14). Wild cluster bootstrap intervals were 0.98 to 1.15 by district and birth year and 0.99 to 1.15 by division and birth year. With the 11 birth years as clusters, the interval was 0.97 to 1.13 (p=0.13; S4 Table). Analytic errors with only 11 clusters gave 1.00 to 1.13 but tend to understate uncertainty. District-specific linear trends gave 1.07 (0.98 to 1.16), and excluding imputed dates gave 1.04 (0.96 to 1.13) (S4 Table). All 1,039 neonatal deaths had age at death recorded in days, and 9 occurred on days 28 to 30. Defining neonatal death as death on days 0 to 27 gave the same estimate (S4 Table).

The mean third trimester temperature anomaly had a ratio of 1.18 per °C (1.02 to 1.38; S4 Table). The anomaly correlated weakly with extreme heat day counts (Spearman 0.24; S2 Table). In one model with both, the anomaly ratio was 1.17 (0.98 to 1.41) and the extreme heat day ratio was 1.01 (0.92 to 1.11). When the anomaly was split into its cooler-than-usual and warmer-than-usual parts, the two parts had similar ratios (1.15, 0.84 to 1.58, and 1.22, 0.91 to 1.62; S4 Table).

## Discussion

In this national study of 46,752 births in Bangladesh, neonatal mortality was highest among births in the cool season, not among births after hot pregnancies. Comparing births within the same place, season and year, estimated prenatal extreme heat was not clearly associated with neonatal death. Per 5 extreme heat days in the third trimester, the data were compatible with anything from a 2% decrease to a 15% increase in risk. Larger contrasts, such as 10 or more days, were estimated imprecisely.

Wang and colleagues conducted the largest study of heat in pregnancy and neonatal death. They used Demographic and Health Survey data from 33 African countries with cluster, birth-year and grid by birth-month fixed effects, a design close to ours [10]. They found 1 to 4 extra neonatal deaths per 1,000 births for 50 extra extreme heat days. Scaled to 50 days, our whole pregnancy risk difference of 0.8 per 1,000 per 5 days is 8 per 1,000 (95% CI −4 to 20). The interval includes their range, but our point estimate is larger. Only 33 births in our data had 50 or more extreme heat days, and the definitions of an extreme day differ, so this comparison is rough. In India, consecutive sweltering days in pregnancy were associated with low birthweight and preterm birth [27]. In South Asia, prenatal humid heat was associated with child stunting [28]. These outcomes are more common than neonatal death, so associations are easier to detect. A 31-country study reported higher neonatal mortality with prenatal heat [11]. Its models included region but no calendar month of birth. A district-level panel study in Ghana linked higher maximum temperature, with lags up to 3 months, to higher neonatal mortality [29]. Its monthly district counts cannot separate prenatal heat from heat after birth. Our design comparison shows why this matters in Bangladesh. Without season control, warmer third trimesters were associated with lower mortality. Births in December to February, whose third trimesters fall mostly in the cooler months, had the highest mortality. Births whose third trimesters fall in warm months were born in seasons of lower mortality, so warmth appeared protective. Where the high-mortality season follows hot months, the same bias could instead create a false harmful association. We cannot say whether this bias affected earlier studies, because their data and seasons differ from ours.

The seasonal pattern itself deserves attention, but our data describe it rather than explain it. Births in December to February, the coolest months, had the highest mortality. A pooled analysis that included Bangladesh found that cold accounted for about twice as many neonatal deaths as heat [3]. In rural Bangladesh, perinatal and respiratory deaths rose at low temperatures, with no clear heat effect [14]. In contrast, in two rural surveillance sites, very severe disease in newborns was more frequent at higher temperatures and in the warmer months [30]. Maternal nutrition, access to care and other seasonal factors could also explain the pattern. A separate analysis of cold after birth would be needed to test a cold effect.

The two temperature metrics measure different things. Extreme heat days count only the hottest 5% of days, while the anomaly averages all departures from the usual temperature. The anomaly result suggests a modest gradient of risk with general warmth across the temperature range, rather than a signal concentrated in the hottest days. However, the anomaly was one of many checks, and its interval included 1 when both metrics were in the same model. A 2026 systematic review of heat thresholds for pregnancy outcomes found the evidence limited by heterogeneity and methodological variability between studies [31]. We do not treat either metric as superior; both point to small or no associations.

Apart from a true absence of effect, several reasons may explain why we found no clear association. First, exposure was measured with error. The two reanalyses agreed only moderately on which days were extreme, and cluster locations were displaced. Such error may attenuate associations, but it can also distort them when it is spatially and seasonally structured. Agreement between the two products shows that results do not depend on the choice of product, not that exposure is accurate, and it cannot remove errors the products share. Second, with about 1,000 deaths, precision was limited, so the study could detect only moderate or larger associations. Some estimates pointed towards harm, such as the ratio for 10 or more third trimester extreme heat days. Because we examined many windows and checks, we treat these as hypotheses to test in larger datasets, not as findings.

Third, heat may act mainly through stillbirth and early pregnancy loss [5,6]. Birth histories record only live births, so heat-related losses before birth would be missing from our sample. If the frailest pregnancies were lost before birth, surviving live births could appear unaffected. The estimate among the poorest households was higher, but the evidence for a difference between wealth groups was weak and the analysis was exploratory. Earlier studies have reported greater vulnerability among poorer mothers [5,10].

For policy, these results do not show that heat in pregnancy is harmless. Heat remains linked to preterm birth and stillbirth [5,6], and mechanisms in pregnancy are plausible [7,8]. Few heat health action plans address pregnant women and newborns [32]. Heat action plans in Bangladesh can still include advice for pregnant women without waiting for neonatal mortality evidence. For surveillance, two changes would help. Birth registration or facility records with gestational age would allow direct study of preterm birth. Releasing exact birth dates with cluster locations, as MICS 2025 did, would let future surveys test these links across more years and settings.

### Strengths and limitations

The study used a national sample with day of birth and cluster locations, which allowed exposure windows of a few weeks. Fixed effects for place, season and year controlled for fixed differences between districts, for seasonality and for national trends. The negative control did not show a clear association, but this cannot rule out residual confounding. It also does not address misclassification of exposure in the true windows. Two reanalysis products and two reference periods gave similar estimates.

The study also has limitations. First, gestational age was not recorded, so windows assumed a 40-week pregnancy. For preterm births, the third trimester window started too early and included days from the second trimester. Because preterm babies die more often, this misclassification may differ between survivors and deaths, and its direction is uncertain. Alternative windows, including an assumed 259-day pregnancy, gave similar results, but we did not formally bound the misclassification. Second, temperature came from coarse reanalyses that agreed only moderately on extreme days. A 0.5° grid cannot capture local heat, such as heat inside homes. Third, the fixed effects do not remove changes over time within districts, such as improvements in care, although district-specific trends did not change the estimate. Fourth, residence was known only at interview; restricting to mothers who had not moved gave a similar result. Fifth, only live births were included, so heat-related stillbirths could not be studied. If heat ended the most vulnerable pregnancies before birth, this would tend to understate any association with neonatal death. Sixth, births are observed only through women who were alive, ever married and resident in the household at the 2025 interview. Births to mothers who died or emigrated are missing, more so for older births. Seventh, wealth and mother’s education were measured at the interview, up to 10 years after birth, which limits the wealth analysis and the control for education. Eighth, we did not examine housing materials, electricity or access to cooling as modifiers of heat risk. Finally, births up to 10 years before the survey carry recall error in dates. Results were similar for births in the last 5 years and after excluding imputed dates.

### Conclusions

In this national birth-history analysis, neonatal mortality varied by season of birth and was highest among births in December to February. When births were compared within the same district, season and year, estimated prenatal extreme heat was not clearly associated with neonatal death. The intervals did not exclude modest increases, and uncertainty in gestational age and exposure may have affected the estimates. Studies of prenatal heat and newborn survival should control for season of birth and improve measurement of gestational age and exposure.

## Data Availability

MICS 2025 data sets are available from UNICEF on registration at mics.unicef.org. The displaced cluster coordinates are released by UNICEF to registered users on request. ERA5 data are available through the Copernicus Climate Data Store and Open-Meteo, and MERRA-2 data through NASA POWER (power.larc.nasa.gov).

https://mics.unicef.org/surveys

## Data availability

MICS 2025 data sets are available from UNICEF on registration at mics.unicef.org. The displaced cluster coordinates are released by UNICEF to registered users on request. ERA5 data are available through the Copernicus Climate Data Store and Open-Meteo, and MERRA-2 data through NASA POWER (power.larc.nasa.gov). The model specification is given in S1 Text. The birth-level file linked to temperature cannot be shared, because it contains the displaced cluster locations that UNICEF releases only to registered users. The numerical values underlying every figure and table are in S1 Data.

## Author contributions

SMAUH conceived the study and designed the methodology. He curated the data, performed the formal analysis, and created the visualizations. He also wrote the original draft, revised the manuscript, and managed the project. MKI contributed to interpretation of the results and reviewed the manuscript. MKH contributed to data curation, wrote part of the original draft, and reviewed and edited the manuscript. MMS and SMRH contributed to interpretation of the results and reviewed the manuscript. KUB supervised the study, validated the results, and reviewed and edited the manuscript. All authors read and approved the final manuscript.

## Supporting information

**S1 Text. Model specification.**

**S1 Fig.**
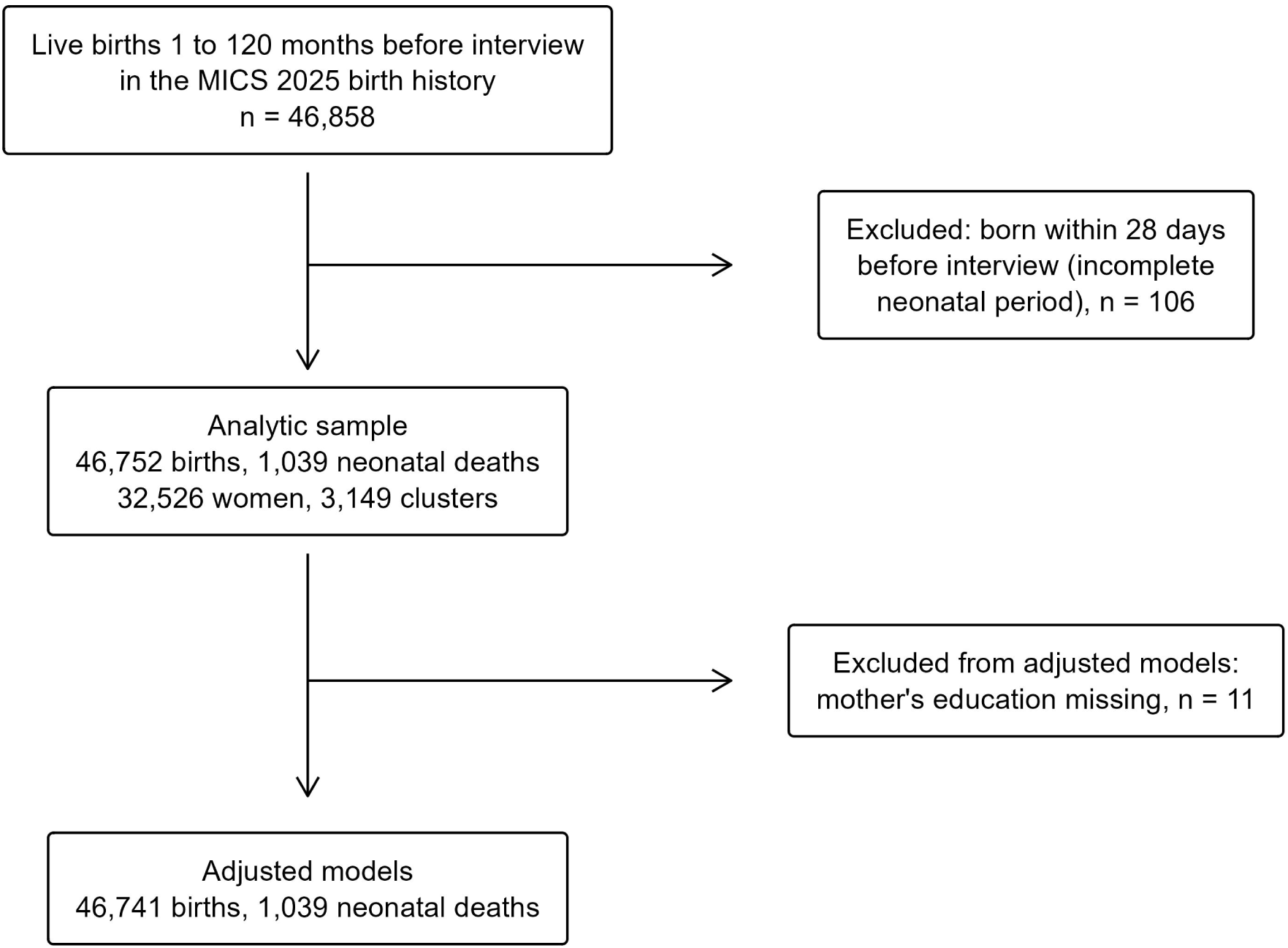
Flow of births into the analysis.

**S2 Fig.**
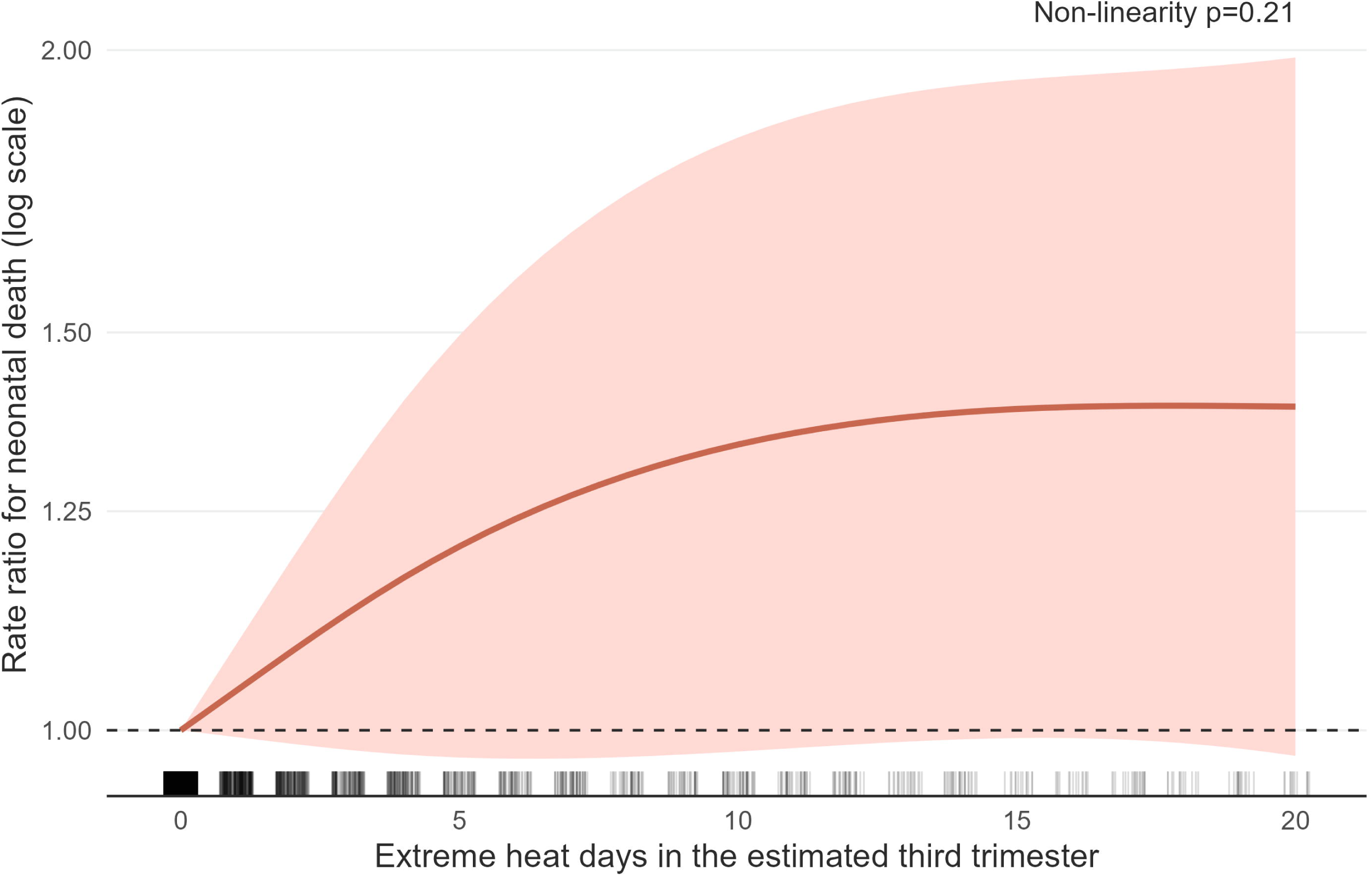
Third trimester extreme heat days and neonatal death, restricted cubic spline.

**S1 Table. Agreement of ERA5 and MERRA-2 temperature, reference periods, and main estimates with each product.**

**S2 Table. Extreme heat exposure in pregnancy: distribution, trend by year of birth and correlations between windows.**

**S3 Table. Estimates under fewer and more fixed effects.**

**S4 Table. Third trimester categories and further checks: thresholds, anomaly, fixed effects, outcomes, imputed dates, calendar windows, variance estimators, clustering, bootstrap and residence.**

**S1 Data. Numerical values underlying the figures and tables.**

